# Cost Savings Associated with Extended Battery Longevity in Cardiac Resynchronization Therapy Defibrillators (CRT-D)

**DOI:** 10.1101/2024.04.17.24306001

**Authors:** Jeffrey L. Williams, Ryoko Sato, Caroline M. Jacobsen

## Abstract

**Background:** Cardiac resynchronization therapy-defibrillators (CRT-D) are devices established as treatment for symptomatic heart failure patients with an indication for CRT and at risk of sudden cardiac death. However, battery depletion poses a significant clinical and economic burden; extended service life may reduce costs due to generator changes and associated complications. The purpose of this study was to estimate the potential cost-savings associated with extended battery longevity in a Medicare patient population receiving CRT-D implantation.

**Methods:** A decision tree was used to explore three battery capacities, which represent the leading device manufacturers available in the US: 1.0 ampere-hours (Ah), 1.6Ah, and 2.1Ah. Yearly risk of all-cause mortality, device-related complications, and end of battery life were estimated. Over a time horizon of 6 years, estimated costs included device implantation, replacement, follow-up appointments, and complications. Costs were discounted at 3%. Univariate deterministic sensitivity analysis was completed for patient survival, battery survival, complication incidence and costs, procedure costs, and time horizon.

**Results:** In the base-case, the average total costs to Medicare over 6 years were $41,527, $48,515, and $56,647 per person (USD 2023) for the 2.1Ah, 1.6Ah, and 1.0Ah devices, respectively. The total per-person replacement cost for the 1.0Ah devices was more than 4 times that of the 2.1Ah devices ($20,126 versus $5,006). When extrapolated to the total number of CRT-D implants each year over a 6-year period, the difference in costs between the extended (2.1Ah) and lowest (1.0Ah) battery capacity exceeded $500 million.

**Conclusions:** Extended longevity CRT-D batteries demonstrate significant cost savings to Medicare over 6 years compared to those with lower battery capacity. These data indicate long-term economic considerations should be included in device selection.

## Introduction

Cardiac resynchronization therapy defibrillators (CRT-D) are an established treatment for a subset of symptomatic heart failure patients at risk for sudden cardiac death.^1^ The service life of CRT-D poses a significant clinical and economic burden^2^ and prolonged device service life is much more important than smaller generator size.^3^ Extended defibrillator battery longevity is preferred by patients^4^ and is more cost-effective for health systems.^5,6^ Battery capacity as measured in ampere-hours (Ah) is the strongest predictor of CRT-D battery longevity. Prior research has reported that CRT-D extended battery life exceeded patient survival in a typical heart failure cohort with reduced ejection fraction.^7^ Extended longevity CRT-D devices not only outlast average patient life expectancy, they also avoid costs of generator changes and associated complications.

CRT-D generator replacement procedures have elevated risks compared to initial implantation, therefore, avoiding additional procedures is a reasonable goal.^8,9^ Implantable cardioverter defibrillator (ICD) replacements are associated with an increased risk for pocket-related surgical re-interventions, and the need for surgical re-intervention increases with every consecutive device replacement.^10^ One in four patients who undergo two or more replacements of cardiovascular implantable electronic device (CIED) develop infection.^11^ Additionally, further hospitalization is associated with increased incidence of adverse events. In a random sample of hospital admissions in Massachusetts in 2018, at least one adverse event was found in nearly one in four cases and approximately one-fourth of such adverse events were preventable.^12^ Recent data reported a 244% increase in cost when three CRT-D generator implant/replacement procedures versus only one were performed among 15,002 Medicare patients who underwent CRT-D implant or replacement from 2009 to 2020.^6^ Given the increased risk of complications and additional costs associated with generator replacement, the objective of this study was to estimate the potential cost-savings associated with extended battery longevity in a cohort of Medicare patients receiving an initial CRT-D implantation.

## Methods

### Model Structure and Assumptions

A Microsoft Excel^®^-based economic model was developed in the form of a decision tree to explore the potential cost-savings associated with increased battery longevity. The model was used to estimate the average costs associated with an initial CRT-D implantation and replacements per person over a 6-year follow-up from a Medicare perspective using the model structure developed by Gadler et al.^13^ This model explored different battery longevities corresponding to three capacities to represent the leading device manufacturers available: 1.0Ah, 1.6Ah, and 2.1Ah. The annual risk of all-cause death,^14,15^ device-related complications,^16^ and the end of battery life^17^ (based on the manufacturer’s longevity estimate) were applied. The 6-year follow-up was based on the real-world experience from a high-volume implanting institution.^17^ Costs for CRT-D implantation, replacement, and follow-up appointments were calculated using the 100% Medicare Standard Analytical Files (SAF) and the national 2023 Medicare payment level for specific diagnosis-related groups (DRG), Ambulatory Payment Classifications (APC), and Current Procedural Terminology (CPT^®^) codes. The frequency of follow-up visits was based on the recommendation from Heart Rhythm Society (HRS).^18^ The costs related to CRT-D-associated complications were obtained from Schmier et al. (2017),^16^ who used Medicare claims to calculate cost. The conditions and procedures used by Schmier et al. to identify complications are described in the Supplemental Material and costs were converted to 2023 United States Dollars (USD) using the consumer price index.^19^ The costs over the 6-year follow-up period were discounted at a rate of 3% and total cost was calculated as the sum of these costs.

The assumptions applied to patients and procedures are presented in Table 1. In the base case, a patient entering the model is indicated for and undergoes implantation of a CRT-D device at the start of the model (i.e., in year 0) and is followed for 6 years. Patient survival probability is the same regardless of device choice. The model assumed device survival would be 100% for the year of implant/replacement and the following year. As a result, a maximum of three replacements for an individual patient could be performed over the model time horizon of 6 years, and in that worst-case scenario, replacement procedures would occur in years 2, 4, and 6.

**Table 1.** Model Assumptions.

| Component | Assumptions |
| --- | --- |
| Patient | • A patient is implanted a CRT-D device as per DRG/APC code at the start of the model (year 0) and is followed for 6 years.<br>• Patient survival is the same regardless of device choice.<br>• The reduction in the patient survival from year 5 to year 6 is similar to the reduction rate in the previous year (sensitivity analysis). Physician guidance was provided for this assumption. |
| Procedure | • The device survival for all manufacturers is 100% at the year of implantation (at year 0) and the subsequent year (year 1);<br>• Sensitivity analysis extends the follow-up period to 10 years. The reduction in the battery survival from year 7 to 10 for 2.1Ah follows the similar reduction rate for that of 1.0 Ah, based on Alam et al. (2007). Physician guidance was provided for this assumption.<br>• There are no replacements due to device malfunctions, only due to battery depletion. |

APC, ambulatory payment classification; CRT-D, cardiac resynchronization therapy defibrillator; DRG, diagnosis-related group

In addition to a base-case analysis, a univariate deterministic sensitivity analysis was performed varying patient survival, battery survival, incidence and costs of complications, procedure costs, and the time horizon.

### Base-Case Analysis and Inputs

The input data used for the base case analysis are shown in Table 2 and include patient survival, battery survival, incidence and costs of complications, Medicare costs of CRT-D implantation and replacement, and the number and cost of follow-up visits. Annual patient survival was obtained from Yao et al. (2007)^14^ who performed a Markov-based Monte Carlo simulation to estimate costs associated with CRT-D therapy from a United Kingdom (UK) healthcare perspective. Event-free battery survival rates were obtained from Alam et al. (2017)^17^ who examined battery longevity of 621 CRT-D recipients at their institution. The source of incidence rates and costs of complications was the simulation by Schmier et al. (2017),^16^ in which both upper and lower bounds were reported (lower bounds were used for the base case). The 100% Medicare SAFs from 2019 to 2021 were used to calculate cost inputs for CRT-D procedures and follow-up visits, using appropriate DRG/APC/CPT^®^ codes and the Medicare 2023 reimbursement amount for each code. Final model inputs represent a weighted average reimbursement for that procedure or visit, where weights reflect the relative volume of claims during 2019-2021 for each DRG, APC, or CPT^®^ code. Details of these calculations can be found in the Supplemental Material.

**Table 2.** Base-Case Input Parameters.

| Parameter | 2.1Ah | 1.0Ah | 1.6Ah | Notes |
| --- | --- | --- | --- | --- |
| Patient survival |  |  |  | Source: Yao et al. (2007) |
| Year 0 | 100% | 100% | 100% |  |
| Year 1 | 95% | 95% | 95% |  |
| Year 2 | 90% | 90% | 90% |  |
| Year 3 | 85% | 85% | 85% |  |
| Year 4 | 81% | 81% | 81% |  |
| Year 5 | 77% | 77% | 77% |  |
| Year 6 | 72% | 72% | 72% |  |
| Event-free battery survival |  |  |  | Source: Alam et al. (2017) |
| Year 0 | 100% | 100% | 100% |  |
| Year 1 | 100% | 100% | 100% |  |
| Year 2 | 98% | 99% | 100% |  |
| Year 3 | 98% | 92% | 100% |  |
| Year 4 | 95% | 74% | 90% |  |
| Year 5 | 90% | 36% | 69% |  |
| Year 6 | 77% | 10% | 44% |  |
| Incidence of complication |  |  |  | Source: Schmier et al. (2017) lower bound |
| Complication first year after primary implant | 4% | 4% | 4% |  |
| Complication first year after replacement | 2% | 2% | 2% |  |
| Infection first year after primary implant | 2% | 2% | 2% |  |
| Infection first year after replacement | 3% | 3% | 3% |  |
| Complication cost to Medicare |  |  |  | Source: Schmier et al. (2017) lower bound; 2023 USD |
| Complication | \$1,112 | \$1,112 | \$1,112 | |
| Infection | \$29,550 | \$29,550 | \$29,550 | |
| Medicare costs of CRT-D |  |  |  | Sources: implantation and replacement costs: claims data (see appendix), reflect |
| Initial implantation | \$34,436 | \$34,436 | \$34,436 | |
| Replacement | \$32,123 | \$32,123 | \$32,123 | |
| Follow-up visit (per visit; 4 visits per year) | \$82 | \$82 | \$82 | weighted average of IP and OP costs using 2023 Medicare reimbursement amounts; visits: frequency from Wikoff et al. (2008), cost reflects weighted average of follow-up visit at facility using 2023 Medicare reimbursement amount |

CRT-D, cardiac resynchronization therapy defibrillator; USD, US dollars; IP, inpatient; OP, outpatient

These inputs were used to calculate the average cost per patient over a 6-year period for initial implantation, replacements, and follow-up visits. These costs were summed to arrive at the total cost per patient to Medicare. To calculate the total cumulative cost of replacements among the CRT-D Medicare population, costs were summed for annual cohorts of patients who were assumed to have received initial implantations during years 0 through 6. Specifically, it was assumed that each year there were 15,577 initial implantations (which represents the average number of annual CRT-D implantations observed in the 100% Medicare SAF claims files during 2019-2021), and each annual cohort had between 0 and 6 years of follow-up costs, depending on the year they entered the model. That is, those who entered at year 0 had 6 years of follow-up costs, those who entered at year 1 had 5 years of follow-up costs, etc. Therefore, the total cumulative cost reflects 109,039 patients who received initial implantations between years 0 and 6 and had between 0 and 6 years of follow-up costs.

### Sensitivity Analysis and Inputs

To perform univariate deterministic sensitivity analysis, individual inputs were varied and the model was re-run to produce alternative cases. For one scenario, patient survival inputs were based on a prospective study (ALTITUDE) of patients who received ICD or CRT-D devices (Saxon et al. 2010).^15^ These data provide estimates for up to 5 years of follow-up; the survival probability for year 6 was derived using the trend from these estimates along with physician input. In a second scenario, the follow-up time was extended to 10 years using similar methods: deriving years 6 through 10 based on base-case battery and patient survival properties during years 0 through 5 and physician guidance. A third scenario explored the effect of using the upper bounds of complication incidence and cost estimates from Schmier et al.^16^ Other scenarios varied the cost to Medicare for CRT-D implantation and replacement to 20% more or 20% less of the base-case amount. The inputs used for these scenarios are shown in Table 3.

**Table 3.**
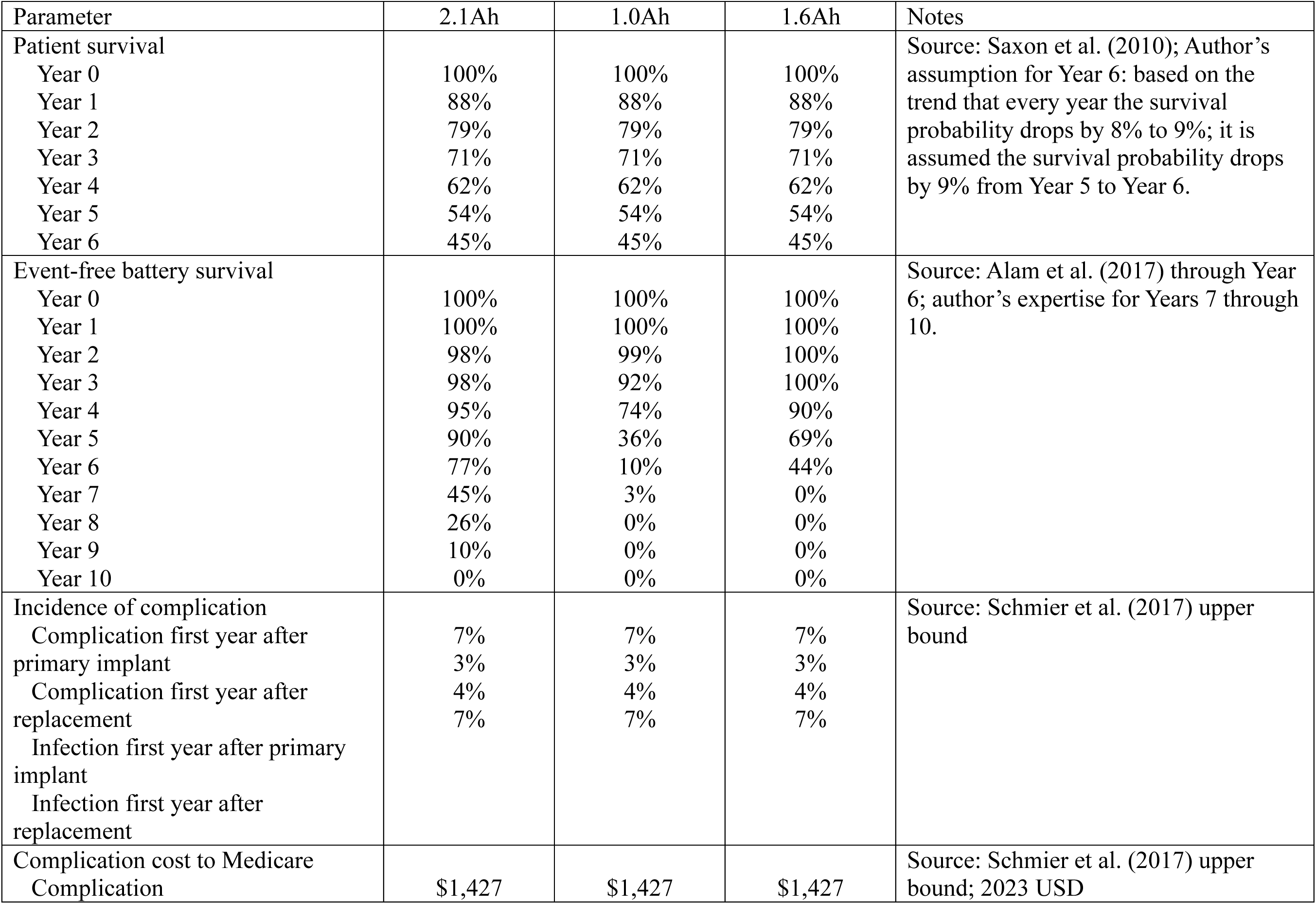
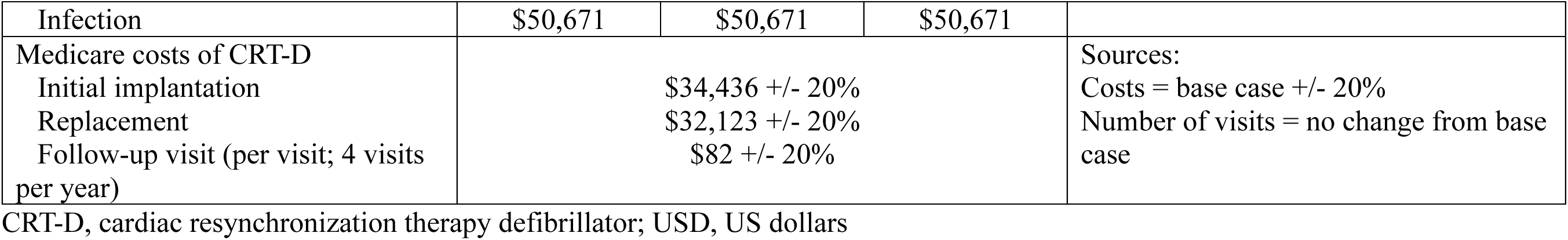
Sensitivity Analysis Input Parameters.

## Results

### Base Case

The average total costs to Medicare associated with a 2.1Ah CRT-D device implantation over 6 years was $41,527 per person (Table 4) in the base-case. The corresponding costs for the 1.6Ah and 1.0Ah devices were $48,515 and $56,647 per person, respectively. The use of a 2.1Ah CRT-D device saved Medicare an average of $15,120 per person compared with the use of a 1.0Ah CRT-D device, and an average of $6,988 per person compared with a 1.6Ah CRT-D device. The costs of the initial implantation, related complications, and routine follow-up visits were the same across devices, thus, the differences in total average per person costs were driven by costs associated with replacements. The total replacement cost for 1.0Ah CRT-D devices ($20,126 per person) was more than 4 times that of 2.1Ah devices ($5,006); the total replacement cost for 1.6Ah CRT-D devices ($11,994) was more than double that of 2.1Ah devices.

**Table 4.** Base-Case Analysis Results.

|  | Average Per-patient Medicare Costs over 6-year Follow-up |  |  |  |  |
| --- | --- | --- | --- | --- | --- |
| Cost Category | 2.1Ah | 1.0Ah | 1.6Ah | Difference between 1.0Ah and 2.1Ah, USD (%) | Difference between 1.6Ah and 2.1Ah, USD (%) |
| Initial Implant |  |  |  |  |  |
| Procedure | \$34,436 | \$34,436 | \$34,436 | No difference by definition | No difference by definition |
| Complications | \$635 | \$635 | \$635 | | |
| Post-procedure | \$82 | \$82 | \$82 | | |
| follow-ups | \$35,154 | \$35,154 | \$35,154 | | |
| Total |  |  |  |  |  |
| Replacements |  |  |  |  |  |
| Procedures | \$4,856 | \$19,524 | \$11,635 | \$14,668 (302%) | \$6,779 (140%) |
| Complications | \$137 | \$552 | \$329 | \$415 (302%) | \$192 (140%) |
| Post-procedure | \$12 | \$50 | \$30 | \$38 (302%) | \$17 (140%) |
| follow-ups | \$5,006 | \$20,126 | \$11,994 | \$15,120 (302%) | \$6,988 (140%) |
| Total |  |  |  |  |  |
| Routine follow-ups | \$1,367 | \$1,367 | \$1,367 | No difference by definition | No difference by definition |
| Total | \$41,527 | \$56,647 | \$48,515 | \$15,120 (36%) | \$6,988 (17%) |
| Cumulative cost of replacement in full Medicare population over 6 years if 15,577 CRT-D implants per year | \$152,681,105 | \$679,635,453 | \$333,012,260 | \$526,954,348 (345%) | \$180,331,156 (118%) |

CRT-D, cardiac resynchronization therapy defibrillator; USD, US dollars
All values are in 2023 USD

Using these values and assuming 15,577 CRT-D implants annually over 6 years, the cumulative replacement cost to Medicare would be $152,681,105 for 2.1Ah devices, $679,635,453 for 1.0Ah devices, and $333,012,260 1.6Ah devices (Table 4). The difference in cumulative replacement costs between the 2.1Ah and 1.0Ah devices was $526,954,348, and $180,331,156 between the 2.1Ah and 1.6Ah devices.

### Sensitivity Analysis

The results of the univariate deterministic sensitivity analyses produced average total costs to Medicare associated with 2.1Ah CRT-D device implantation over 6 years per patient that ranged from $34,640 - $48,414 (Table 5 and Figure 1). The highest and lowest total cost estimates occurred when the procedure cost inputs were varied to 20% higher and 20% lower than the base-case values. The per person savings to Medicare for using the 2.1Ah CRT-D device ranged from $10,815-$18,054 compared with the 1.0Ah device, and $4,727-$8,344 compared with the 1.6Ah device.

**Table 5.** Sensitivity Analysis Results.

| Scenario | Average Per-patient Medicare Costs over 6-year Follow-up |  |  | Difference between 1.0Ah and 2.1Ah, USD (%) | Difference between 1.6Ah and 2.1Ah, USD (%) |
| --- | --- | --- | --- | --- | --- |
|  | 2.1Ah | 1.0Ah | 1.6Ah |  |  |
| Scenario 1: Base Case | \$41,527 | \$56,647 | \$48,515 | \$15,120 (36%) | \$6,988 (17%) |
| Scenario 2: Patient survival (Saxon) | \$39,757 | \$50,572 | \$44,483 | \$10,815 (27%) | \$4,727 (12%) |
| Scenario 3: Battery survival (Alam + 10 years of follow-up) | \$55,030 | \$66,715 | \$58,886 | \$11,685 (21%) | \$3,856 (7%) |
| Scenario 4a: Procedure costs -20% | \$34,640 | \$49,760 | \$41,627 | \$15,120 (44%) | \$6,988 (20%) |
| Scenario 4b: Procedure costs +20% | \$48,414 | \$63,534 | \$55,402 | \$15,120 (31%) | \$6,988 (14%) |
| Scenario 5a: Replacement costs -20% | \$40,555 | \$52,742 | \$46,188 | \$12,187 (30%) | \$5,632 (14%) |
| Scenario 5b: Replacement costs +20% | \$42,498 | \$60,551 | \$50,842 | \$18,054 (42%) | \$8,344 (20%) |
| Scenario 6: Complication rate (Schmier upper bound) | \$42,331 | \$57,996 | \$49,571 | \$15,665 (37%) | \$7,240 (17%) |
| Scenario 7: Complication costs (Schmier upper bound) | \$42,058 | \$57,471 | \$49,181 | \$15,412 (37%) | \$7,123 (17%) |

All values are in 2023 USD

**Figure 1.**
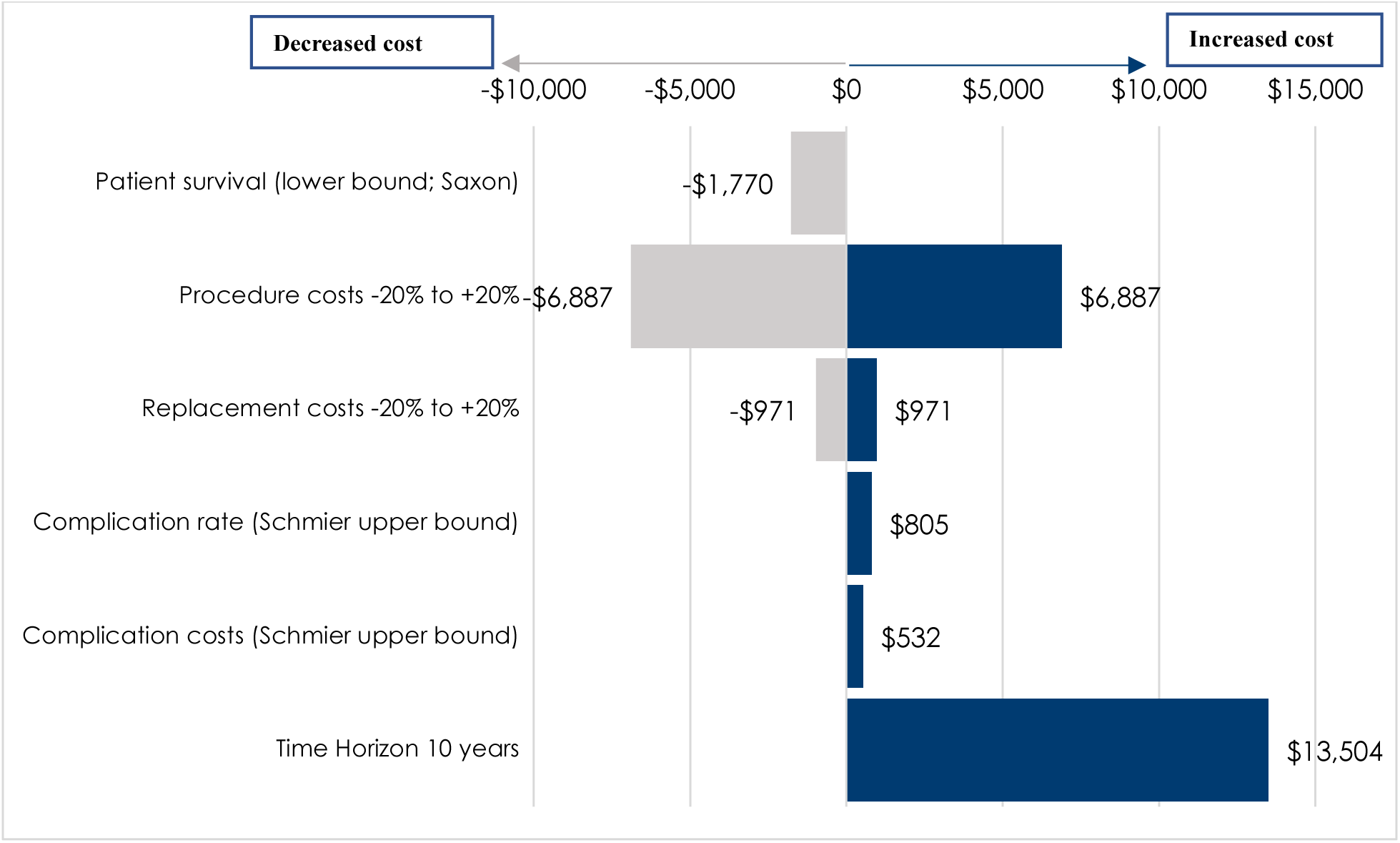
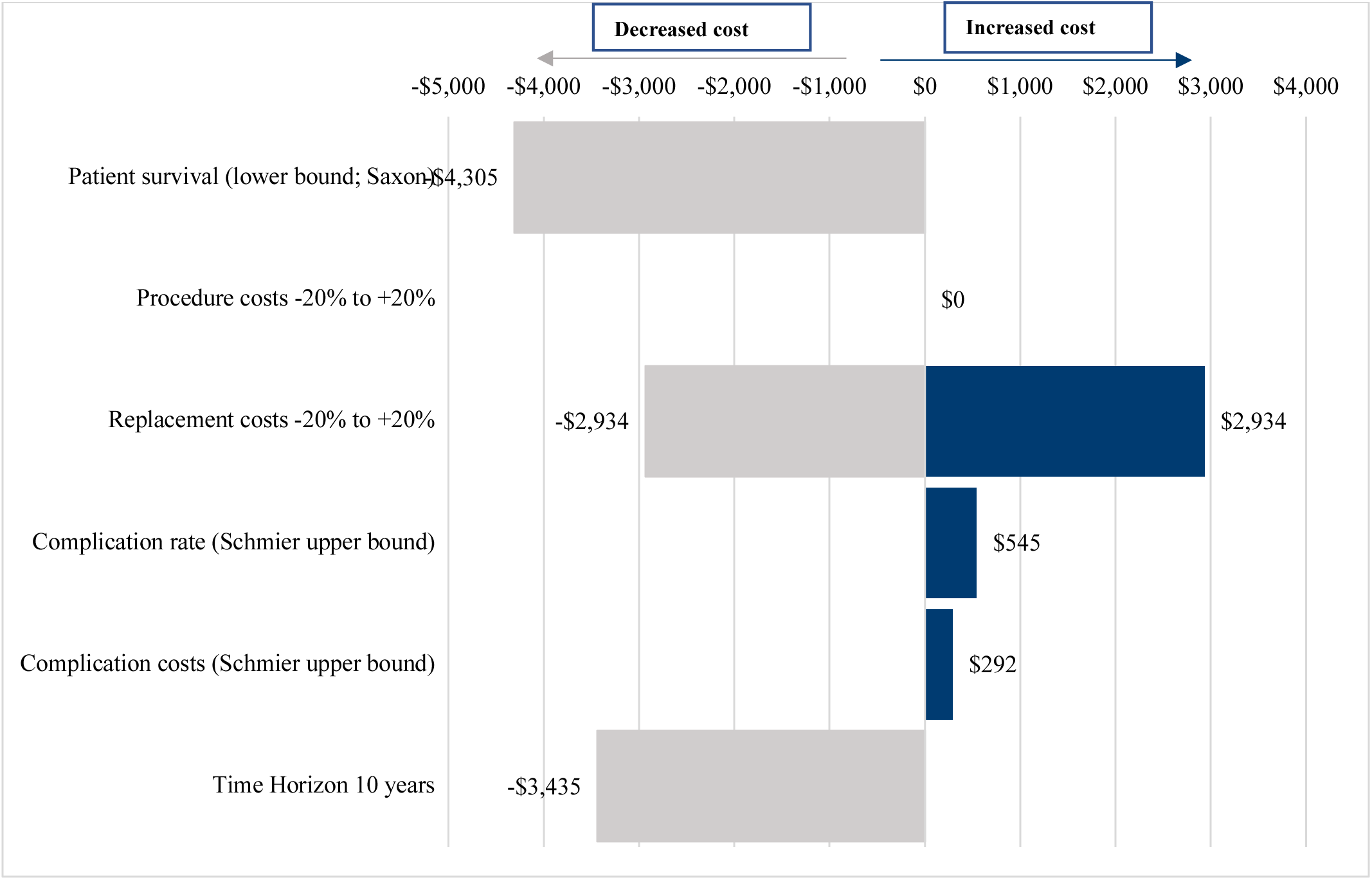
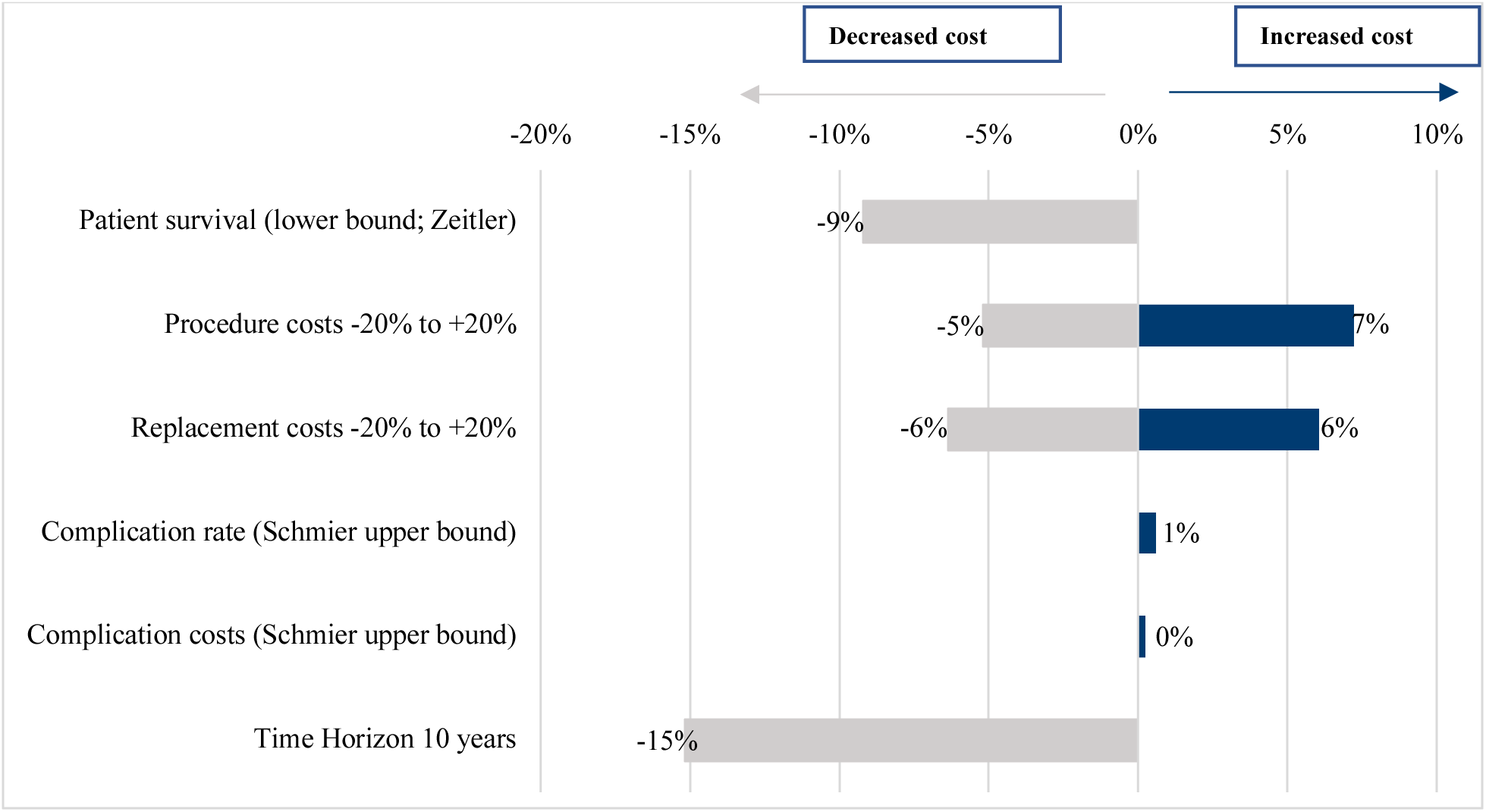
Tornado Diagrams (All values in 2023 USD) for: A. Average Cost per Patient Over 6 Years with 2.1 Ah Battery [Base: 2.1 Ah CRT-D costs $41,523] B. Dollar Savings Associated with 2.1 Ah Battery versus 1.0 Ah Battery [Base: cost-saving $15,118] C. Percent Savings Associated with 2.1 Ah Battery versus 1.0 Ah Battery [Base: 36%]

The average total cost per person to Medicare associate with CRT-D implantation and replacement over a 10-year time horizon was $55,030 (2.1Ah), $58,886 (1.6Ah), and $66,715 (1.0Ah). That value was 7% higher for 1.6Ah devices and 21% higher for 1.0Ah devices compared with 2.1Ah devices (Table 5).

## Discussion

The data presented here demonstrate significant cost savings when extended longevity CRT-D devices are used in Medicare patients. The cumulative cost of replacement in the Medicare cohort over 6 years was $152.7 million for the 2.1Ah device versus $333.0 million and $679.6 million for devices with 1.6Ah and 1.0Ah batteries, respectively. The difference in costs between the 2.1Ah and 1.0Ah devices was over $527.0 million (345% higher).

CRT-D replacement due to battery depletion is a significant cost-driver for payors^13,20^ and a significant complication-driver for patients.^21,22^ Landolina et al. (2017) found the need for device replacements at 6 years was reduced from 83% to 68% with the use of devices with improved battery longevity.^20^ Modeling has shown that increased utilization of extended longevity CRT-D led to a 39% annual reduction in major complications and a 12.8% reduction in total annual costs ($496 million) for Medicare.^23^ A prior study examining 15,002 Medicare patients who underwent CRT-D implant or replacement from 2009 through 2020 reported a total cumulative cost to Medicare for a patient undergoing 1, 2, and 3 generator implant or replacement procedures to be $52,795, $88,976, and $128,846, respectively.^6^ These data demonstrate the substantial increased costs to Medicare when patients are subjected to repeat CRT-D generator changes that extended longevity devices may help to reduce. More importantly, longevity seems to be more important to patients than the size of the device; most prefer a larger device when it is accompanied by greater longevity.^4^ Guidelines should consistently emphasize the importance of patient preferences in all clinical decisions.^24^ The value offered by extended longevity CRT-D led the National Institute for Health and Care Excellence, which provides guidance to the National Health Service (NHS) of the UK, to conclude extended longevity CRT-D benefits patients, are associated with fewer procedures, and save the NHS approximately £6 million within first 5 years of utilization.^5^ The Board of Medicare Trustees determined in 2023^25^ that the Hospital Insurance trust fund is not adequately financed over the next 10 years; the program can only guarantee eight years of paying 100% of scheduled benefits to over 65 million Americans. Incremental improvements to clinical practice that reduce complications and improve cost-effectiveness may help the financial stability of the Medicare program.

While this study has several strengths, the results should be viewed in light of its limitations. First, the available data on input parameters such as patient survival, battery survival, complication rate, infection rate are limited. Some of these data are from older studies, from countries other than the US (the setting for this study), and conditional on other factors such as co-morbidity. Secondly, the model does not include societal costs among patients, which include transportation and opportunity costs. Finally, the model also does not consider patient preference, although previous research suggests patients would prefer options with extended battery longevity.

## Conclusion

The per person and estimated cumulative cost to Medicare for generator replacement of CRT-D devices is substantial. The exclusive use of extended longevity devices over 6 years would save Medicare between $15,120 and $6,988 per person, and between $180 million and $527 million cumulatively. Adequate consideration of the multiple factors affecting device choice, including the economic impact of generator replacement due to battery longevity, should be a consideration in physician decision making at the time of initial implant.

## Data Availability

The data that support the findings of this study are available on request from the corresponding author.

## Non-standard Abbreviations and Acronyms

CRT-D: Cardiac resynchronization therapy defibrillators
Ah: ampere-hours
CIED: of cardiovascular implantable electronic device
ICD: Implantable cardioverter defibrillator
DRG: Diagnosis-related group
APC: Ambulatory Payment Classifications
CPT: Current Procedural Terminology
HRS: Heart Rhythm Society
SAF: Standard Analytical Files
NHS: National Health Service

## Acknowledgements

The authors would like to thank Craig Solid and Ally Pachelli for assistance in preparing this manuscript, as well as Claire Duxbury for her contributions to the model.

## Supplemental Material

List of Complications Used by Schmier et al. (2017)

| Category | Description |
| --- | --- |
| Complications, infection | Septicemia<br>Endocarditis<br>Cellulitis<br>Fever<br>Shock<br>Bacteremia<br>Infection due to device |
| Complications, non-infection | Chest tube insertion<br>Pericardiocentesis<br>Pulmonary embolism<br>Hemopericardium<br>Cardiac tamponade<br>Unspecified disease of pericardium<br>Pleural effusion<br>Pneumothorax<br>Respiratory arrest<br>Hematoma |

<u>Calculation of Costs to Medicare for Implantation and Replacement:</u>

Inpatient Initial Implantation:

**Calculation of Costs to Medicare for Implantation and Replacement:****Inpatient Initial Implantation:**
| DRG Code | Claim Volume (Number), 2019-2021 | Claim Volume (Percent) | Medicare 2023 Reimbursement Amount (USD) | Weighted Average |
| --- | --- | --- | --- | --- |
| 222 | 1,533 | 14% | \$52,520 | \$42,485 |
| 223 | 313 | 3% | \$35,798 | |
| 224 | 1,409 | 13% | \$48,628 | |
| 225 | 708 | 7% | \$34,693 | |
| 226 | 3,900 | 37% | \$43,907 | |
| 227 | 2,731 | 26% | \$34,439 | |

DRG, diagnosis-related group; USD, US dollar

Outpatient Initial Implantation:

| APC Code | Medicare 2023 Reimbursement Amount (USD) |
| --- | --- |
| 5232 | \$32,076 |

APC, ambulatory payment classification

Weighted Average of Inpatient and Outpatient Initial Implantation Costs

| Site of Service | Claim Volume<br>(Number), 2019-2021 | Claim Volume<br>(Percent) | Average<br>Medicare 2023<br>Reimbursement<br>(USD) | Weighted<br>Average |
| --- | --- | --- | --- | --- |
| Inpatient | 10,594 | 22.7% | \$42,485 | \$34,436 |
| Outpatient | 36,136 | 77.3% | \$32,076 | |

Data Source: Centers for Medicare and Medicaid Services. 2019-2021 Medicare 100% standard analytic files. DRG/APC Code list.

Weighted Average of Inpatient and Outpatient Replacement Costs

| Site of Service | Claim Volume<br>(Number), 2019-2021 | Claim Volume<br>(Percent) | Average<br>Medicare 2023<br>Reimbursement<br>(USD) | Weighted<br>Average |
| --- | --- | --- | --- | --- |
| Inpatient (DRG<br>245) | 1,127 | 3.4% | \$33,447 | \$32,123 |
| Outpatient (APC<br>5232) | 31,842 | 96.6% | \$32,076 | |

Weighted Average of Follow-up Visit Costs

| Follow-Up Visit<br>CPT Code | Claim Volume<br>(Number), 2019-2021 | Claim Volume<br>(Percent) | Average<br>Medicare 2023<br>Reimbursement<br>for Facility<br>Location (USD) | Weighted<br>Average |
| --- | --- | --- | --- | --- |
| 93289 | 2,116 | 6.9% | \$73.87 | \$82.30 |
| 93284 | 4,189 | 13.6% | \$107.76 | |
| 99212 | 2,174 | 7.1% | \$35.58 | |
| 99213 | 11,416 | 37.1% | \$66.08 | |
| 99214 | 10,214 | 33.2% | \$97.60 | |
| 99215 | 697 | 2.3% | \$143.34 | |

Data Source: Centers for Medicare and Medicaid Services. 2019-2021 Medicare 100% standard analytic files. DRG/APC Code list.

